# The Story of Shukuru – a film to enhance knowledge, attitudes, and practices regarding schistosomiasis and related elimination interventions in Pemba, Tanzania

**DOI:** 10.64898/2026.10.29.26366827

**Authors:** Lydia Trippler, Ulfat Amour Mohammed, Hasina Suleiman Amour, Adam Nassor Ali, Saleh Juma, Said Mohammed Ali, Stefanie Knopp

## Abstract

**Background:** Schistosomiasis is a neglected tropical disease affecting individuals in 79 countries, primarily in sub-Saharan Africa. The World Health Organization recommends the treatment, snail control, and behavior change interventions to decrease prevalence, but culturally tailored interventions remain rare. Here we aim to determine the impact of an educational film on schistosomiasis-related knowledge, attitudes, and practices among schoolchildren and adults in Pemba, Tanzania.

**Methodology/Principal Findings:** From November 2023 to February 2024, a survey was conducted among adult community members in 20 administrative areas in Pemba on their preferences for a planned film about schistosomiasis. The film was produced in March 2024 based on these suggestions and filmed in Pemba. In July 2024, a pretest-posttest study in ten administrative areas assessed the film’s impact on schistosomiasis-related knowledge, attitudes, and willingness to change behavior among children and adults. Binomial generalized linear mixed-effects models estimated changes in knowledge and attitudes before versus after viewing. Of 882 participants surveyed in 2023/24, 810 (91.8%) were willing to watch a film about schistosomiasis, most commonly on TV (545, 67.3%). The impact study included 195 children and 195 adults. The odds of a correct knowledge answer were 39% higher after viewing (OR 1.39, 95% CI 1.29-1.50, p<0.001), and the odds of a positive attitude were 77% higher (OR 1.77, 95% CI 1.47-2.12, p<0.001). Before the film, 71.9% (200/278) of participants used natural water bodies for daily activities, and 72.5% (145/200) of these never used protective clothing. Afterward, 88.2% (240/272) reported being likely to avoid water bodies and 54.4% (148/272) likely to use protective clothing.

**Conclusions/Significance:** An educational film developed from community input and filmed locally increased knowledge and attitudes among children and adults and fostered willingness to adopt self-protective behaviors. However, willingness alone is insufficient without infrastructure (e.g., wells, tap water) to change behavior. Therefore, such investments remain essential for schistosomiasis elimination.

**Author Summary:** Schistosomiasis is a disease caused by parasitic worms that live in open freshwater bodies, and it affects people in many countries, most heavily in sub-Saharan Africa. People become infected through everyday contact with contaminated water, such as swimming, washing, or farming. The World Health Organization recommends treatment, snail control and behavior change to reduce the disease, but most educational efforts are not tailored to local culture. We worked with communities on the island of Pemba, Tanzania, to create an educational film about schistosomiasis, shaped by residents’ own ideas about what such a film should look like and where they would watch it. We then tested whether the film changed people’s knowledge, attitudes, and stated intentions around water contact, comparing children’s and adults’ responses before and after watching. Knowledge and positive attitudes both improved after viewing, and many participants said they intended to avoid risky water contact and use protective items such as rubber boots. However, good intentions are not enough on their own: without safer alternatives like wells or tap water stations, people may have no real choice but to keep using open water bodies. Eliminating schistosomiasis will require investment in this kind of infrastructure alongside education.

## Background

Human schistosomiasis is a neglected tropical disease (NTD) that currently puts people from 79 countries at risk of infection, primarily in sub-Saharan Africa [1, 2]. Schistosomiasis can cause serious health consequences, including chronic urinary tract fibrosis, anemia, and bladder cancer [3–5]. The primary intervention against schistosomiasis in endemic countries is preventive chemotherapy through mass drug administration (MDA) of praziquantel, implemented by national schistosomiasis control programs [1, 6]. In addition to treatment interventions, the World Health Organization recommends (WHO) implementing snail control and behavior change interventions and the construction of safe water and hygiene infrastructure in endemic areas [6].

High coverage and compliance with these interventions are essential for their sustained success and for progress toward the WHO’s goal of eliminating schistosomiasis as a public health problem globally by 2030 [1, 7, 8]. However, achieving and maintaining high intervention compliance remains an ongoing challenge in many endemic settings.

Behavior change communication interventions in the context of schistosomiasis aim to enable healthy behaviors and reduce risky behaviors related to schistosomiasis infection and transmission, thereby sustaining the gains achieved by treatment-based control programs and accelerating progress toward elimination [9].

On the Zanzibar islands, a combination of MDA, snail control, and behavior change interventions led to the elimination of schistosomiasis as a public health problem in 2017, and the islands now aim to interrupt *Schistosoma haematobium* transmission [10–12].

Behavior change communication tools used on the islands have included a comic strip booklet for schoolchildren [13, 14] and, more recently, community movie nights, play days in schools and the construction of washing platforms developed through a human-centered design approach during the Zanzibar Elimination of Schistosomiasis Transmission (ZEST) studies [15] and implemented during the ZEST and SchistoBreak studies [10, 16–18]. However, compliance with MDA among adults in Zanzibar remains low [19], and existing behavior change communication tools have primarily engaged children, leaving a gap for similarly engaging tools aimed at adults.

Here we aim to present the full procedure undertaken before and after the production of an educational film about schistosomiasis by i) assessing adult community members’ preferences regarding the content and format of a film about schistosomiasis, ii) creating a film about schistosomiasis based on the communities’ suggestions, and iii) determining the impact of the film on schistosomiasis-related knowledge, attitudes, and practices in schoolchildren and adult community members in Pemba, Tanzania.

## Methods

### Study setting

The study was conducted on Pemba Island, which along with Unguja Island and many small islands, constitutes the Zanzibar archipelago, a semi-autonomous region of the United Republic of Tanzania. The Zanzibar archipelago is located approximately 30 km off the eastern coast of mainland Tanzania. Pemba is divided into four districts, which are subdivided into 129 small administrative areas, called shehias [20]. The formative survey was implemented in 20 shehias that constituted the study sites of the SchistoBreak study, which was conducted from 2020 to 2024 in the two districts Wete and Micheweni in the north of Pemba [21]. Based on the most recent population census, approximately 95,000 people lived in these 20 shehias in 2022 [20]. The film was shot in different locations in these two districts. The impact evaluation study was conducted in ten of the 20 SchistoBreak shehias.

### Study design and participants

The presented study was divided into three parts: i) Conducting a questionnaire survey among community members in Pemba to gather their suggestions regarding the content and format of a potential educational film about schistosomiasis from November 2023 to February 2024, ii) Shooting a movie that follows the ideas raised by the community in March 2024, iii) Assessing the impact of the final movie on knowledge, attitudes, and practices regarding schistosomiasis among schoolchildren and community members in Pemba (impact study) in July 2024.

Participants were interviewed about their suggestions regarding the content and length of a potential film about schistosomiasis and related interventions. The survey questions were embedded in a larger questionnaire deployed as part of the 2023/2024 cross-sectional community-based survey of the SchistoBreak study in the north of Pemba [21]. Due to the study design, only adults were eligible to participate in the questionnaire. Any individual aged ≥18 was eligible to participate, provided that they had given written informed consent.

Once the survey was completed, a film crew was hired to shoot an educational film in March 2024, following the story of a boy who tests positive for schistosomiasis (see *Film description* below for details). Once the film was shot and edited, it was shown to communities and schools on a big screen.

To assess the impact of the film on knowledge, attitudes and practices (KAP) of schoolchildren and community members, questionnaires following a pretest-posttest design were conducted with participants before and after they watched the film.

### Data collection for the formative film-development survey

The cross-sectional SchistoBreak survey was conducted from November 2023 to February 2024 in 20 small administrative areas in the north of Pemba Island. For the survey, 80 housing structures per administrative area were pre-randomized and identified by the local enumerators through a navigation system paired with a questionnaire application, both installed on Samsung Galaxy Tab A tablets. Details on the randomization and identification of households are presented elsewhere [22]. One adult household member was invited to participate in the questionnaire survey in each identified household. Participants were asked for their suggestions regarding the content, length, and format of a potential educational film about schistosomiasis.

### Filmmaking

Following the community’s input from the questionnaire survey on what kind of film they would like to watch and what they would like to learn about urogenital schistosomiasis, a storyboard was created in preparation for the filmmaking, and a Tanzanian film crew (J&O Communications, http://jandocommunications.co.tz/) was hired. Furthermore, during the film shooting in March 2024, two employees of the Zanzibar Ministry of Information were always present. The actors in the film were local community members and local study team members. The film was shot in Kiswahili, however, in an edited version, English subtitles were added.

### Data collection for impact study

In July 2024, the film was shown in ten shehias. In each shehia, the film was presented in the biggest primary school and in a public area of the shehia community.

In each school, 21 children from grade five were randomly selected to participate based on the sample size calculation (see Sample size calculation). In case the school had multiple classes in grade five, the largest class according to the school board was selected. In the class, the children were asked to line up according to their sex, and the number of children in each group was counted. Afterward, the total number of children per group was divided by the desired group size (ten or eleven), and every nth child in the lineup was selected based on the division result. Subsequently, the study aim and procedure were explained to the children. Furthermore, demographic details of the selected children were registered, and consent forms for the parents were handed out. In addition to the written consent by the parents or legal guardians, children aged twelve years and older were asked to sign an assent form. On the following day, after returning the written consent from their parents, the children were randomly assigned into three groups using a computer-generated randomization list. Children in groups one and two were invited to participate in a questionnaire interview about knowledge, attitudes, and practices regarding schistosomiasis. Subsequently, all the children watched the film. Following the film screening, children in groups one and three were asked to participate in a similar questionnaire (again) regarding schistosomiasis.

In each community, the sheha (head of shehia) had selected one location to conduct the study, as it was paramount to have electricity access in order to show the film using a projector. Furthermore, in advance of the arrival of the study team, the shehas had informed their communities about the study and invited as many people as possible from the area to join the screening. In the communities, only adult individuals were selected to participate, following the same procedure as in the schools. Participants were asked to line up by sex and every nth participant was selected and then randomly assigned to one of three groups. However, if the number of community members present was fewer than or exactly the required number, which was 21, all available individuals were included in the study, following a convenience sampling approach, without regard to sex. Community members in groups one and two were invited to participate in a questionnaire interview about knowledge, attitudes, and practices regarding schistosomiasis. Subsequently, all the community members watched the film. Following the film, individuals in groups one and three were asked to participate in a similar questionnaire (again) regarding schistosomiasis.

### Sample size calculation

A sample size calculation was run to assess the number of participants needed to assess the impact of the film on KAP scores regarding schistosomiasis. Adjusting for a 95% confidence interval, 80% statistical power, a small effect size, and a 20% drop-out rate, the results revealed that 420 individuals should be invited to participate to include a minimum of 336 individuals in the final analysis, corresponding to 210 individuals invited (168 minimum in the final analysis) in both the schools and communities.

### Statistical Methods

The data cleaning and analyses were performed with R version 4.4.3.

To assess knowledge and attitudes, scores were calculated in the following way: Each correct answer to a knowledge item was scored 1 and each incorrect answer was scored 0. For the attitude sentences, a positive attitude was scored 1 and a negative attitude was scored 0. The same scoring was applied to the before- and after-film questionnaires. The median of these proportions from all participants was calculated for the before and after questionnaires, respectively. Results are reported as median (range). The analyses for knowledge and attitudes were divided into three comparisons i) all participants together (groups 1-3), ii) only participants who have answered questions before AND after the film watching (group 1 only), and iii) participants who have either only answered questions before or after watching the film but not both (groups 2 and 3).

To assess the association between film and knowledge and attitudes, a binomial generalized linear mixed model (GLMM) was fitted separately for knowledge and attitudes, for each of the three comparisons mentioned above. The outcome was the number of items a participant scored 1 on, out of the total number of items covered. Explanatory variables were demographic variables, such as age and sex, and the assessment time point (before or after watching the film). Occupation and interaction terms were included in selected models and compared against simpler models using likelihood ratio tests. The implementation unit (school/community) was included as a random effect in every model. Participant identity was additionally included as a random effect for comparisons i) and ii), which include repeated measurements from the same individuals, and an observation-level random effect was included throughout to account for residual overdispersion.

For practices, individuals’ answers to practices regarding schistosomiasis were summarized in bar charts. Because the post-film questionnaire was administered immediately after the film, participants had not yet had the opportunity to change their actual behavior. The post-film practice questions therefore asked about intended future behavior, on a scale from “very unlikely” to “very likely,” rather than current behavior, and for this reason the before- and after-film responses were presented separately in bar charts rather than compared directly as a before/after change. Participants’ responses on whether they liked the film, and any further comments, were also summarized.

### Data management

All data collected during the questionnaire interviews in parts one and three of the study were collected using the application Open Data Kit (ODK, www.opendatakit.org) installed on Samsung Galaxy Tab A tablets. All data collected with ODK were sent to and stored on a secure server of the Swiss Tropical and Public Health Institute, Allschwil, Switzerland. Furthermore, no data that could potentially identify an individual will be shared with third parties. Signed informed consent forms are stored for ten years at the Public Health Laboratory – Ivo de Carneri in Pemba, United Republic of Tanzania.

### Ethics approval and consent to participate

The study protocol for the formative survey as part of the SchistoBreak study was waived by the ethics committee in Switzerland (Ethikkommission Nordwest- und Zentralschweiz; EKNZ) on October 23, 2019 (Req-2019-00951). It subsequently received annual ethical approval by the Zanzibar Health Research Institute (ZAHRI). The first approval was given on December 13, 2019 (ZAHREC/03/PR/December/2019/12), and the latest approval was given on March 31, 2023 (ZAHREC/04/AMEND/MARCH/2023/03). The study was registered prospectively at ISRCTN (ISRCTN91431493).

The study protocol for the impact study was approved by the ethics committee in Zanzibar on May 30, 2024 (ZAHREC/01/PR/MAY/2024/18) and waived by the ethics committee in Switzerland (Ethikkommission Nordwest- und Zentralschweiz; EKNZ) on June 24, 2024 (AO_2024-00063).

At the onset of the formative film-development survey and the impact study, respectively, a meeting was held with the community leaders and school principals of the study area to explain the research aims, and study and intervention procedures. Community leaders and school principals were requested to inform their communities and parents of the schoolchildren about the forthcoming schistosomiasis activities. In the meeting preceding the impact study, the film was also shown to the community leaders and principals.

All individuals participating in the study were informed in detail about the objectives and procedures of the study by the local research team and a leaflet that was provided together with informed consent forms. Individuals who were invited to participate in the study could ask questions to the study team. Moreover, the information leaflet included the telephone number of the local principal investigator so that participants or parents of participating children could call in case they had additional questions. Study participants provided written informed consent for their participation. For participating children aged <18 years, written consent was obtained from their parents or legal guardians. Children aged 12-17 years additionally signed an assent form themselves. In the 10 study schools, any child enrolled in grade five was eligible to participate, provided that the parents had given written informed consent and children aged 12 years and older had additionally provided written assent. In the 10 communities, any individual aged ≥18 was eligible to participate, provided that they had given written informed consent.

## Results

### Participation and preferences in the formative film-development survey

In the SchistoBreak household-based questionnaire survey 2023/2024, a total of 882 individuals were asked about their suggestions regarding the content and format of a potential film. Of those, 715 (81.1%) were women and 167 (18.9%) were men. The median age was 43 (18-86).

Of the 882 participants, 810 (91.8%) indicated a willingness to watch a film about schistosomiasis (Figure 1A). Of those, 691 (85.3%) indicated that they would be amenable to watching a film of 15 minutes in length or longer (Figure 1B). A total of 448 (55.3%) individuals were interested in watching every topic related to schistosomiasis (Figure 1C). Another 357 (44.1%) were particularly interested in the transmission, symptoms, complications, prevention, and/or treatment of schistosomiasis and/or the parasite itself. Three (0.4%) individuals had other wishes, such as *“We should be provided with education about other diseases in order to achieve better protection”*. Only two (0.2%) individuals had no specific wishes for a topic to be watched in the movie. A total of 545 (67.3%) respondents preferred to watch the film on TV, and 312 (38.5%) were also open to watching it during a community meeting (Figure 1D). A total of 49 (6.0%) individuals indicated that they would watch the film via WhatsApp and 38 (4.7%) via YouTube on their smartphones, while eight (1.0%) indicated they would watch it on TV while travelling on a ferry (Figure 1D). A total of ten (1.2%) individuals did not have a preferred place to watch the movie, and another nine (1.1%) individuals mentioned other places, such as *“School”* or *“Everywhere”*.

**Figure 1.**
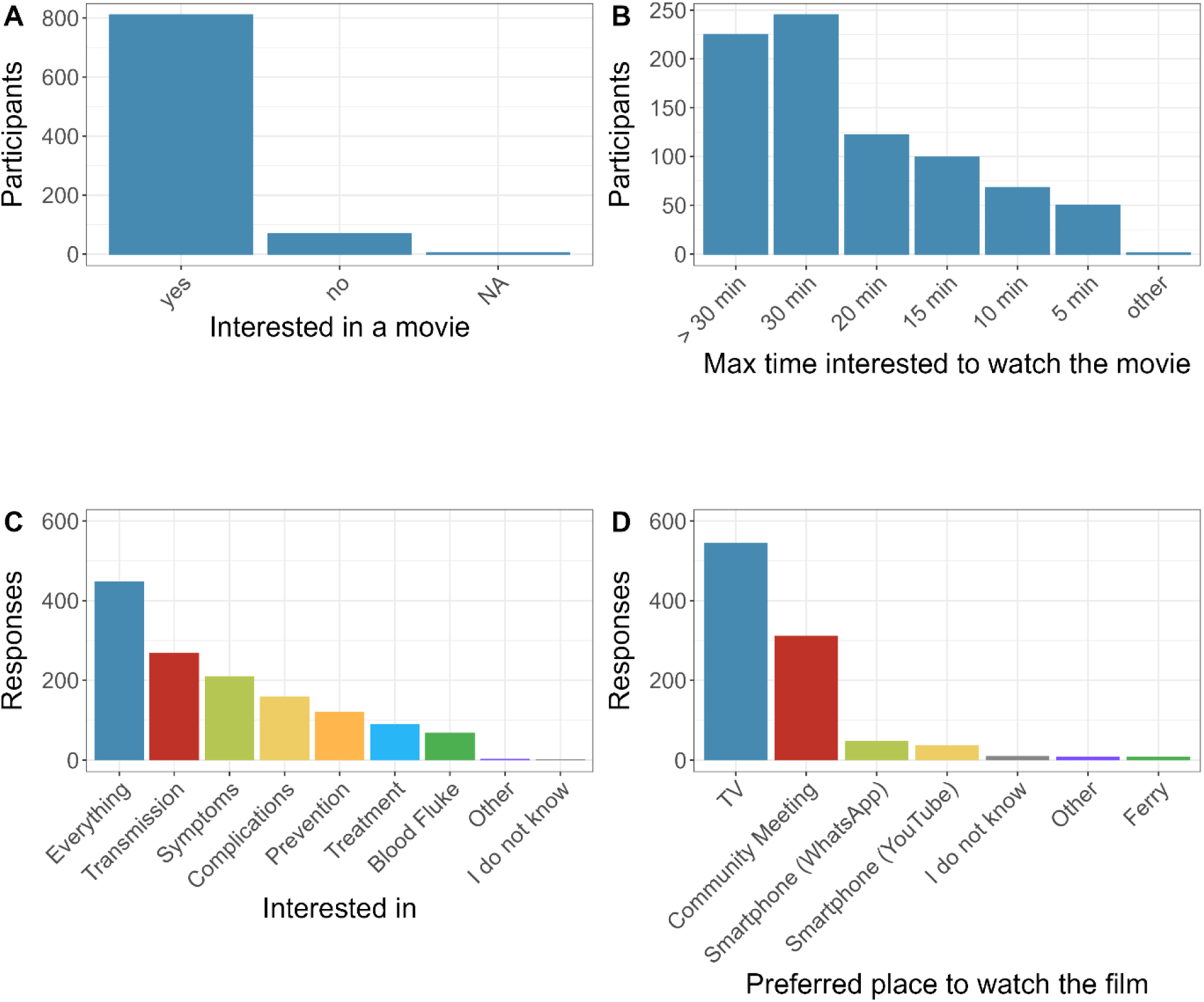
Community members’ preferences about making a movie about schistosomiasis (A: N=882; B: N=810). Questions C and D allowed multiple responses.

### The film: “The story of Shukuru”

The shooting and editing of the film resulted in an 11-minute-long movie that narrates the story of a boy named Shukuru. The beginning of the film depicts Shukuru, a boy from Pemba, playing and swimming in a natural open freshwater body. Additionally, his mother is shown washing clothes at the same water body. Weeks later, a research team visits Shukuru’s school and tests the children for urogenital schistosomiasis. Shukuru submits a sample of bloody urine.

The urine samples of the children are transported to a laboratory for analysis, where they are filtered and examined for the presence of *Schistosoma* eggs using a microscope. Upon returning to the school, the research team treats Shukuru and other children who tested positive for schistosomiasis with praziquantel. Additionally, the film shows the study team conducting a school outreach day at Shukuru’s school with games and drama to educate the children about schistosomiasis, including the life cycle, preventive measures, and symptoms. By the end of the film, Shukuru returns home from school and tells his mother about the necessary precautions to avoid future infections. The full movie, including English subtitles, can be streamed on https://m.youtube.com/watch?v=OjvuZfVFCis&feature=youtu.be.

### Participation in the study on the impact of the film on knowledge, attitudes, and practices

In the schools, 210 children were registered on the first day of the study. Of these, 11 (5.2%) did not return on the second day, and two (1.0%) returned without signed consent forms from their parents. Thus, 197 children were eligible to participate in the study. Of these, two children did not return to answer the questionnaire after watching the film. The 195 remaining children were randomly assigned to one of three groups: Group 1 (n = 78, 40.0%), Group 2 (n = 59, 30.3%), and Group 3 (n = 58, 29.7%). In the communities, 210 individuals were registered before watching the film. Of these, 14 (6.7%) individuals from groups 1 or 3 did not return to answer the questionnaire after watching the film. Furthermore, one (0.5%) individual had two different data entries for the questionnaire after the film. As the data lacked sufficient clarity regarding the correct entry, this person was excluded from the study. In total, 195 individuals were included in the analysis, of whom 79 (40.5%) were assigned to Group 1, 62 (31.8%) to Group 2, and 54 (27.7%) to Group 3 (Figure 2).

**Figure 2.**
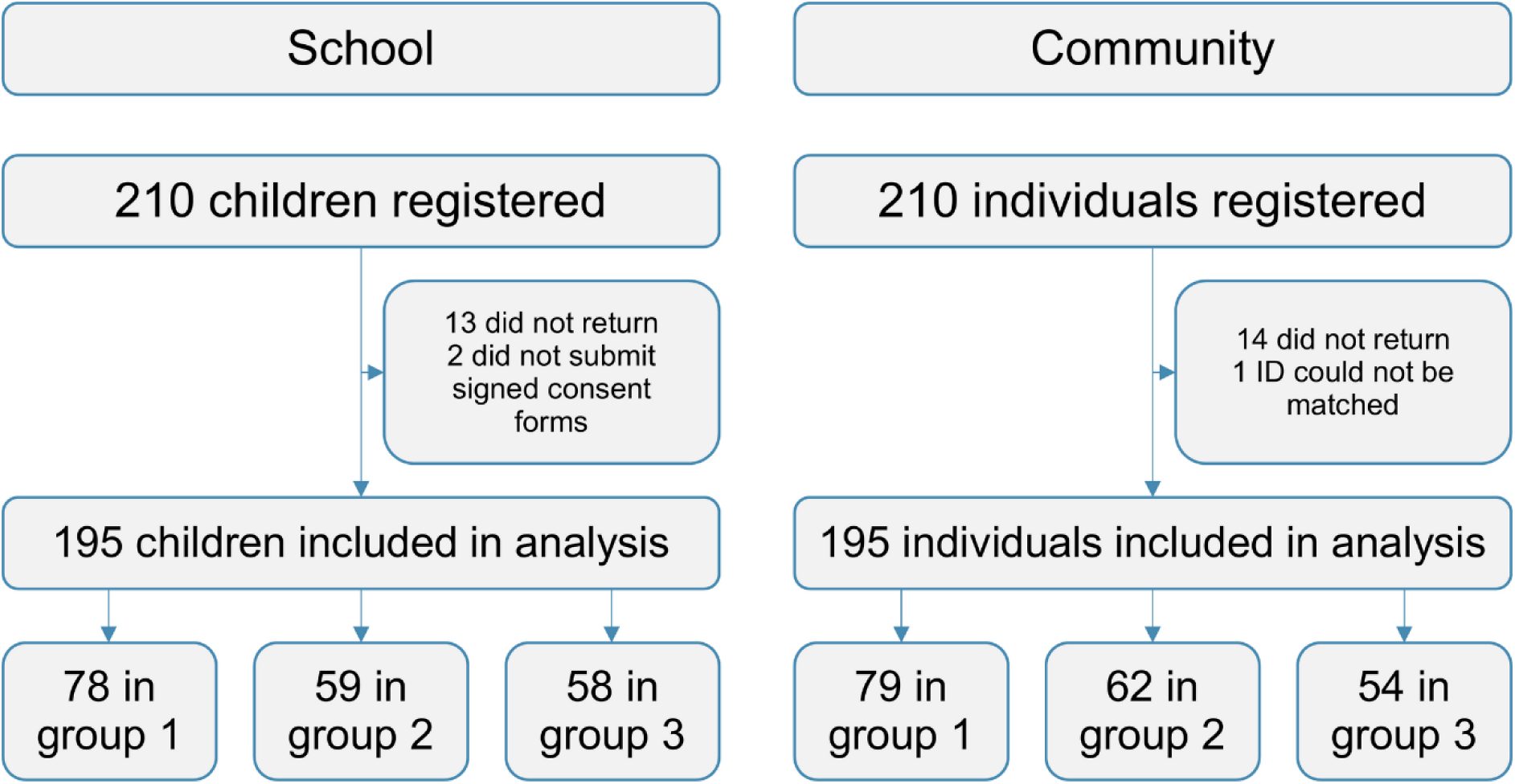
Flowchart of participants in the study to assess the impact of the film on knowledge, attitudes, and practice in schoolchildren and community members. Group 1 = participants answered the questions before and after watching the film; Group 2 = participants answered the questions before watching the film only; Group 3 = participants answered the questions after watching the film only.

### Characteristics of the participants of the impact study

Of the 195 schoolchildren, 98 (50.3%) were female, and 97 (49.7%) were male. The median age was 12 with ranges from 10 to 15 years. Of the 195 community members, 123 (63.1%) were female, and 72 (36.9%) were male. The median age was 40 with ranges from 18 to 82 years. The majority of the community members were farmers (79/195, 40.5%) or indicated not having an occupation (38/195, 19.5%) (Table 1).

**Table 1.** Characteristics of the participants of the impact study in schools and communities.

|  | **Total** | **Schools** | **Communities** |
|---|---|---|---|
|  | (N=390) | (N=195) | (N=195) |
| **Sex** |  |  |  |
| Female | 221 (56.7%) | 98 (50.3%) | 123 (63.1%) |
| Male | 169 (43.3%) | 97 (49.7%) | 72 (36.9%) |
| **Age (years)** |  |  |  |
| Median [Min, Max] | 16.5 [10.0, 82.0] | 12.0 [10.0, 15.0] | 40.0 [18.0, 82.0] |
| **Occupation** |  |  |  |
| Student | 197 (50.5%) | 195 (100%) | 2 (1.0%) |
| Farmer | 79 (20.3%) | 0 (0%) | 79 (40.5%) |
| No work | 38 (9.7%) | 0 (0%) | 38 (19.5%) |
| Business | 17 (4.4%) | 0 (0%) | 17 (8.7%) |
| Fisherman | 14 (3.6%) | 0 (0%) | 14 (7.2%) |
| Teacher | 14 (3.6%) | 0 (0%) | 14 (7.2%) |
| Fundi | 13 (3.3%) | 0 (0%) | 13 (6.7%) |
| Other | 9 (2.3%) | 0 (0%) | 9 (4.6%) |
| Health worker | 3 (0.8%) | 0 (0%) | 3 (1.5%) |
| Housewife | 3 (0.8%) | 0 (0%) | 3 (1.5%) |
| Driver | 2 (0.5%) | 0 (0%) | 2 (1.0%) |
| Ministry | 1 (0.3%) | 0 (0%) | 1 (0.5%) |

### Impact of the film on schistosomiasis-related knowledge

Among all study participants, the median knowledge score was 0.32 (0-0.6) before watching the film and 0.4 (0.08-0.64) after watching the film (Figure 3A). The mixed-effects model indicated that the odds of a correct answer were 39% higher after watching the film than before (OR = 1.39, 95% CI 1.29-1.50, z = 8.8, p < 0.001). Among children, the odds of a correct answer were 43% higher after the film (OR = 1.43, 95% CI 1.28-1.59, z = 6.0, p < 0.001). Among adults, the odds were 35% higher after the film (OR = 1.35, 95% CI 1.23-1.50, z = 6.0, p < 0.001). Among the participants who answered questions before and after watching the film (group 1), the median knowledge score was 0.32 (0-0.56) before watching the film and 0.4 (0.12-0.56) after watching the film (Figure 3B). The mixed-effects model indicated that the odds of a correct answer were 37% higher after watching the film (OR = 1.37, 95% CI 1.25-1.50, z = 6.5, p < 0.001). Among children, the odds were 45% higher after the film (OR = 1.45, 95% CI 1.27-1.66, z = 5.4, p < 0.001). Among adults, the odds were 29% higher after the film (OR = 1.29, 95% CI 1.14-1.47, z = 3.9, p < 0.001).

**Figure 3.**
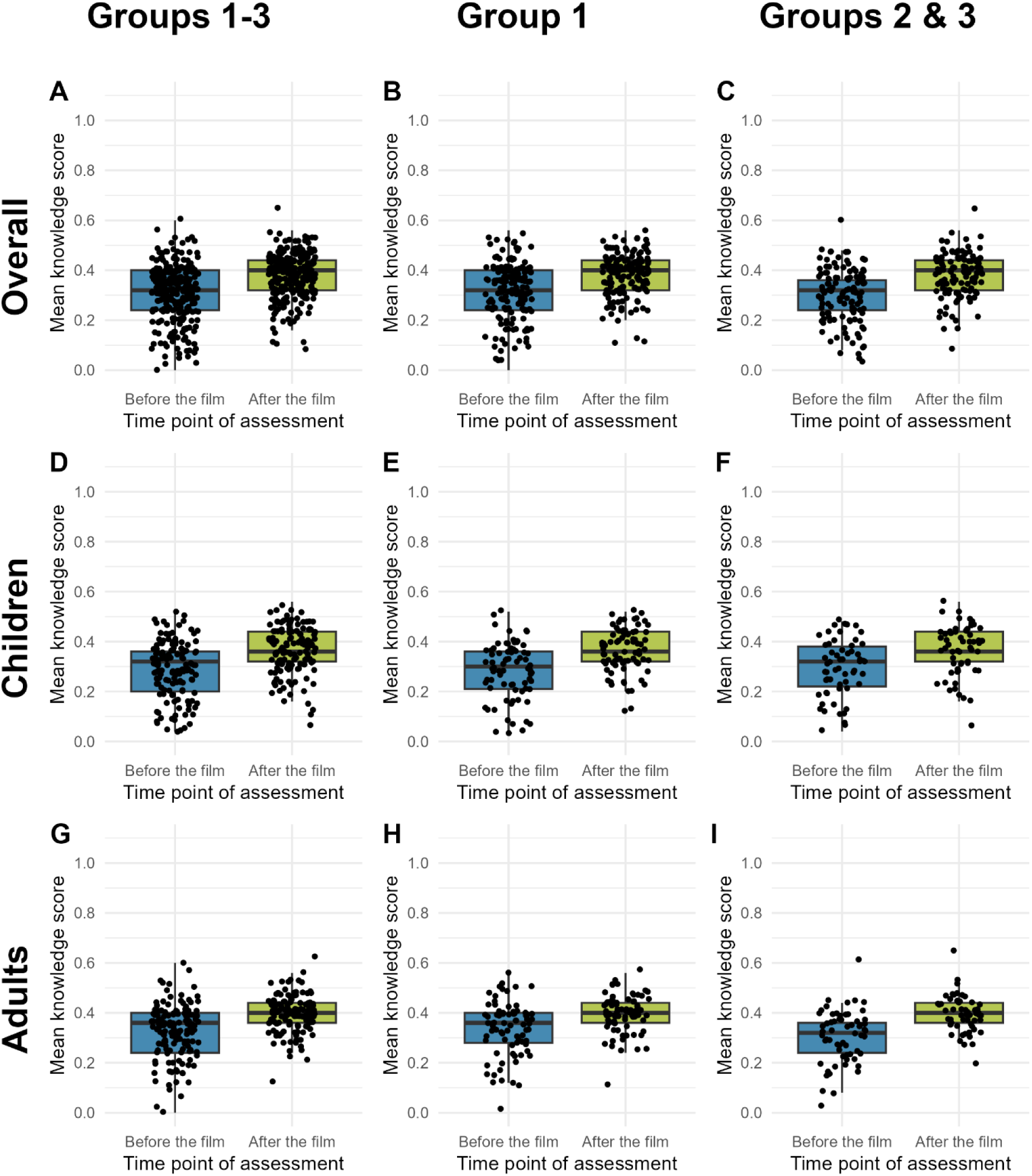
Knowledge scores before and after watching the film among all participants (A, D, G), among participants who answered questions before and after the film (group 1) (B, E, H), and among participants who answered the questions before the film only (group 2) or after the film only (group 3) (C, F, I), shown separately for the overall population (A-C), schoolchildren (D-F), and community members (G-I). In panels C, F, and I, the “before” boxes are group 2 (who only answered before the film) and the “after” boxes are group 3 (who only answered after the film). These are different people shown side by side for comparison, not the same individuals measured at two time points, unlike panels A/B, D/E, and G/H.

### Impact of the film on schistosomiasis-related attitude

Among all study participants, the median attitude score was 0.8 (0.1-1) before and 0.9 (0.1-1) after watching the film (Figure 4A). The mixed-effects model indicated that the odds of a positive attitude were 77% higher after watching the film (OR = 1.77, 95% CI 1.47-2.12, z = 6.1, p < 0.001). Among children, the odds were 93% higher after the film (OR = 1.93, 95% CI 1.53-2.43, z = 5.5, p < 0.001). Among adults, the odds were 54% higher after the film (OR = 1.54, 95% CI 1.15-2.07, z = 2.9, p = 0.003). Among the participants who answered questions before and after watching the film (group 1), the median attitude score was 0.8 (0.1-1.0) before watching the film and 0.9 (0.1-1.0) after watching the film (Figure 4B). The mixed-effects model indicated that the odds of a positive attitude were 72% higher after watching the film (OR = 1.72, 95% CI 1.37-2.16, z = 4.7, p < 0.001). Among children, the odds were 84% higher after the film (OR = 1.84, 95% CI 1.37-2.46, z = 4.1, p < 0.001). Among adults, the odds were 59% higher after the film (OR = 1.59, 95% CI 1.10-2.30, z = 2.5, p = 0.014).

**Figure 4.**
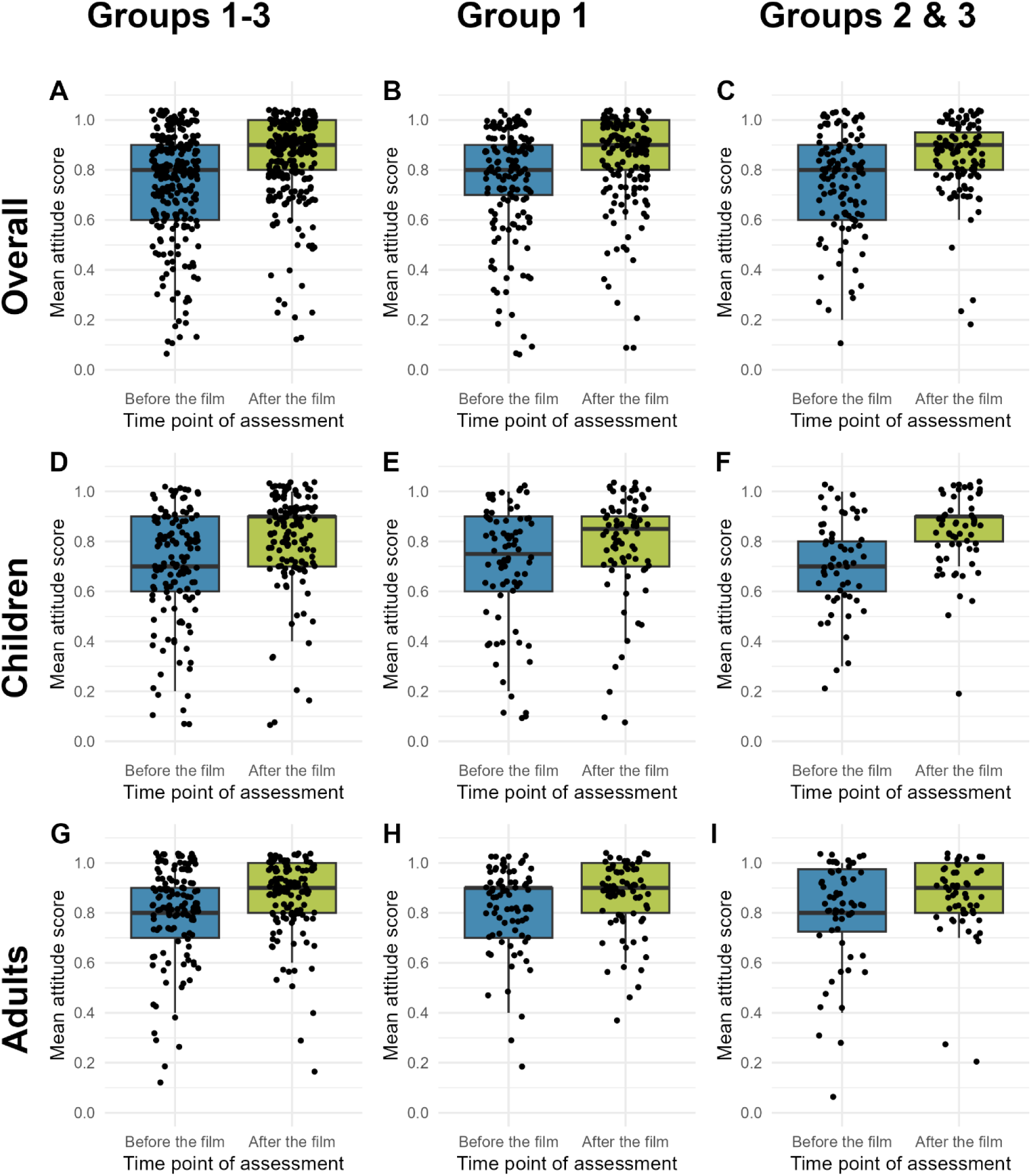
Attitude scores before and after watching the film among all participants (A, D, G), among participants who answered questions before and after the film (group 1) (B, E, H), and among participants who answered the questions before the film only (group 2) or after the film only (group 3) (C, F, I), shown separately for the overall population (A-C), schoolchildren (D-F), and community members (G-I). In panels C, F, and I, the “before” boxes are group 2 (who only answered before the film) and the “after” boxes are group 3 (who only answered after the film), i.e. different people shown side by side for comparison, not the same individuals measured at two time points, unlike panels A/B, D/E, and G/H.

### Impact of the film on schistosomiasis-related practices

Prior to watching the film, 71.9% (200/278) of the participants indicated using water from natural open freshwater bodies for rice farming, swimming, washing clothes, or washing dishes (Figure 5A). Among those, 72.5% (145/200) indicated that they never utilize protective clothing in water bodies or rice fields (Figure 5C). Following the watching of the film, 88.2% (240/272) of the individuals indicated that they were likely or very likely to refrain from utilizing water from natural open freshwater bodies in the future (Figure 5B). Furthermore, 54.4% (148/272) of the individuals indicated that they will likely or very likely utilize protective clothing, such as rainwater boots, in water bodies in the future (Figure 5D). However, another 37.1% (101/272) indicated that they were unlikely or very unlikely to use protective clothing in the future. Before watching the film, 62.6% (174/278) of the participants had already participated in a community meeting about schistosomiasis (Figure 5E). After the film, 93.4% (254/272) said that they would be likely or very likely to participate in future community meetings about schistosomiasis (Figure 5F). Before watching the film, 72.3% (201/278) of the participants had already participated in a MDA (Figure 5G). After the film, 96.0% (261/272) stated that they would likely or very likely participate in future MDAs (Figure 5H).

**Figure 5.**
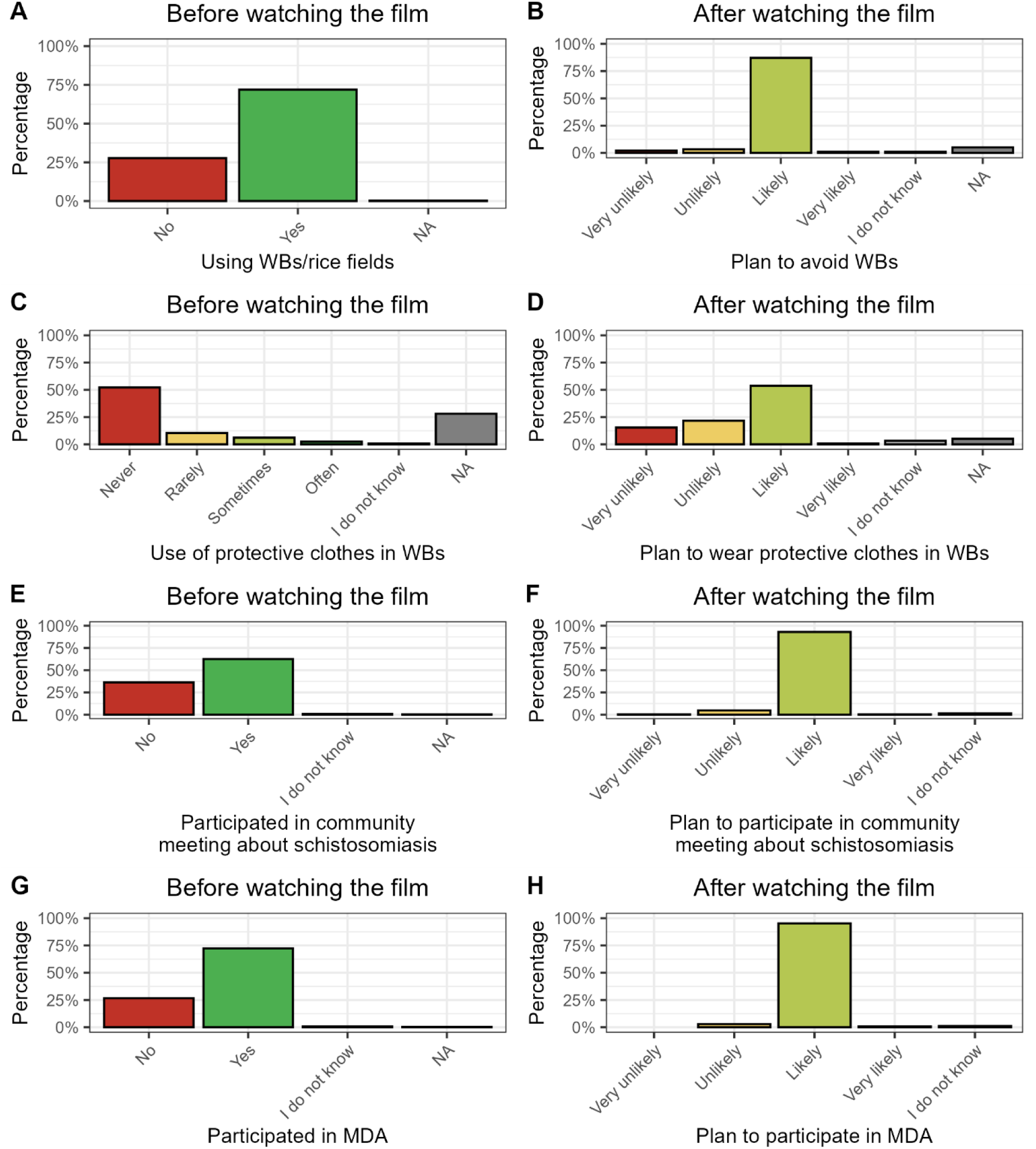
Practices of all participants before watching the film (A, C, E, G) and willingness to change behavior after watching the film (B, D, F, H). WB = water body, MDA = Mass drug administration, NA = Not applicable.

Of the 104 individuals from groups 1 and 3 who were asked about their perception of the film by the end of the questionnaire, 103 (99.0%) individuals indicated that they liked the film. The majority (78/104, 75.0%) of the individuals in groups 1 and 3 did not provide additional comments when asked whether they had anything else to say by the end of the questionnaire interview after watching the film. Two individuals, however, specifically highlighted a lack of infrastructure and financial resources to implement the preventive measures depicted in the film:

*Person 1: “Thank you for educating us about schistosomiasis; we enjoyed and understood the film, but we have no other option but to go to the river because water is scarce for us, and this is due to the lack of wells.”*

*Person 2: “Because we do not have money, we do not have [protective] shoes to wear“ [addition by author]*.

## Discussion

The Zanzibar islands are committed to interrupting *S. haematobium* transmission in the near future. However, fully breaking the *Schistosoma* life cycle and achieving elimination of schistosomiasis on the islands, will require engaging behavior change communication tools that also target the adult population to achieve an increase in knowledge and attitudes and provoke a behavior change.

Our results showed that the schistosomiasis-related knowledge and attitudes of the study participants were significantly higher after watching the movie than before. This was observed regardless of whether participants had been asked questions about schistosomiasis before the film or not, ruling out any learning bias. Furthermore, schistosomiasis-related knowledge was significantly higher after the film not only in the overall study population but also when stratified into children and adults, demonstrating that the intervention was effective in the often-overlooked adult population. Basing the creation of our behavior change communication intervention on community suggestions therefore appeared effective, which is consistent with findings from a study in Mozambique, where community-centered interventions, such as community dialogue increased knowledge and attitudes regarding schistosomiasis among community members [23]. Similarly, a previous study in Zanzibar demonstrated that a comic strip booklet featuring a similar storyline as the film (a boy infected with schistosomiasis) led to increased knowledge in children after one year of implementation in schools [13]. These findings suggest that educational interventions, whether through film or print, can effectively increase knowledge and improve attitudes toward schistosomiasis and related interventions (e.g., participation in MDA and use of protective clothing), especially when adapted to the local culture and informed by community input on preferred content, length, and screen format. Improving knowledge and attitudes is critical for the interruption of *Schistosoma* transmission, as other studies, such as one conducted in Brazil, have shown that higher levels of knowledge correlate with better compliance to interventions, ultimately reducing prevalence [24].

Our results also showed that despite the overall improvements in knowledge and attitudes, participants’ knowledge did not reach a full level of understanding as no individual achieved full knowledge (defined as a score of 1.0, i.e., correctly answering all knowledge items) after watching the film. Hence, a single exposure to the film may not be sufficient for full knowledge acquisition and participants may need to watch the film multiple times to achieve deeper comprehension. Given that the film has already been produced, it can be easily used repeatedly in communities or shared through YouTube and smartphones without significant additional costs. However, during the formative survey, only a small number of individuals mentioned that they would like to watch the film on smartphones. Therefore, we suggest integrating the film into a broad behavioral intervention scheme, where it serves as an engaging component alongside presentations or community dialogues to reinforce knowledge gain and behavior change. We provided the principals of the study schools with copies of the film, so that it can be incorporated into school curriculums to educate children further about schistosomiasis. Furthermore, we suggested that local health authorities, in collaboration with national broadcasters, consider showing the film on public television, e.g. in the halftime of e.g. football games, or during advertisement breaks to reach particularly boys and men who have been the population predominantly infected with *Schistosoma* in Zanzibar and elsewhere [12, 25]. Health facilities could be another important place to reach at-risk people with behavior change activities, e.g. during their waiting time, and to show the film if a computer and screen is available. Targeting health facilities that include a mother and childcare clinic could primarily reach pregnant women or mothers. Preventing *Schistosoma* infections in pregnant women is crucial, as studies have shown that schistosomiasis can contribute to anemia, particularly in cases of severe infections [26, 27]. Anemia in pregnant women is associated with low birth weight and maternal death [28, 29].

However, are increased knowledge and attitudes actually enough to change behavior? While the implementation of control interventions has led to reductions in schistosomiasis prevalence in several countries, including South Africa and Ethiopia [30–32], some countries, such as in Côte d’Ivoire and the Philippines, continue to report low coverage and compliance with interventions, particularly among adults [33, 34]. The results of our study indicated that individuals used the water bodies frequently, and predominantly without protective clothing. Following the film, a significant proportion of respondents indicated a willingness to avoid contact with water bodies in the future or to utilize protective clothing when using water bodies. It is, however, possible that the respondents provided answers that were in line with what they believed we would want to hear. Furthermore, there was no opportunity to observe a change in behavior due to the immediate administration of the questionnaire following the film. Nevertheless, a study conducted in mainland Tanzania, showed that a participatory hygiene and sanitation intervention resulted in a significant decrease in frequency, duration, and timing of water contacts, as reported and observed two years after implementation [35]. However, in some regions, it is difficult for individuals to avoid contact with freshwater bodies, or to use toilets if an inadequate number of latrines are available for use, as shown in a study in Uganda [36]. A study in Brazil showed that compliance with treatment interventions increased significantly in a group that received health education versus a group that did not. However, the self-reported risk behavior remained high despite the health education [24]. In our study, one participant asserted that in the absence of safe water sources, such as wells, there is no alternative but to use water from freshwater bodies. The Ugandan study also demonstrated that it is challenging for community members to refrain from using water bodies, and impossible for those who are fishermen [36]. In Zanzibar, many people are rice farmers, for whom it is impossible to avoid contact with water, which is why they require the use of protective clothing, such as rain boots when going to rice farming. However, in our study, one participant indicated that while they would be amenable to using protective clothing in theory, they lacked the financial resources to procure them (“*Because we do not have money, we do not have [protective] shoes to wear*”). These findings suggest that while knowledge and attitudes may be enhanced through behavior change interventions such as the film, actual behavior change may not occur in the absence of safe water infrastructure or financial resources to purchase protective clothing even if there is a willingness to change behavior.

Overall, our results showed that engaging behavior change tools, such as our film, particularly if created based on community input on content and format, can increase the knowledge and attitudes of schoolchildren and adults. Furthermore, individuals are willing to adopt self-protective behaviors that may reduce their risk of contracting or transmitting *Schistosoma* infections. However, willingness to change behavior alone is not sufficient to achieve the elimination of schistosomiasis if the necessary resources and infrastructure to actually behave differently are not available. Hence, investments in safe water infrastructure, such as wells or tap water stations, are needed to support the elimination of schistosomiasis.

## Data Availability

The minimal dataset and dictionary are attached to the publication as supplementary information.

## Declarations

### Consent for publication

Every person shown in the film provided written informed consent for their identifiable likeness to be included in the film and made publicly available. In the case of children, the parents or legal guardians provided this written informed consent.

### Author contributions

Conceptualization: Lydia Trippler, Said Mohammed Ali, Stefanie Knopp

Data Curation: Lydia Trippler

Formal Analysis: Lydia Trippler

Funding Acquisition: Lydia Trippler, Stefanie Knopp

Investigation: Lydia Trippler, Hasina Suleiman Amour, Adam Nassor Ali

Methodology: Lydia Trippler, Ulfat Amour Mohammed

Project Administration: Lydia Trippler

Resources: Saleh Juma, Said Mohammed Ali

Supervision: Lydia Trippler, Said Mohammed Ali, Stefanie Knopp

Visualization: Lydia Trippler

Writing – Original Draft Preparation: Lydia Trippler

Writing – Review & Editing: Lydia Trippler, Stefanie Knopp

## Acknowledgments

We are very thankful for all schoolchildren and adult participants in our formative and impact study. Thank you for giving us your time to answer questions and watch the film. We also want to thank all the people who have agreed to appear in the film, particularly the actors from the communities and the local study team. Ania Bialic, affiliate public engagement manager at the Glasgow Centre for Parasitology (formerly the Wellcome Centre for Integrative Parasitology), thank you for your advice for storyboard creation, this was fantastic. Layla Hasler from Swiss TPH, thank you for supporting the planning of the filmmaking. And we are especially grateful to J&O Communications, particularly to James, Ami and Godlisten for creating and shooting the film with us. Lastly, we would like to especially appreciate Ibrahim Said Ali for his role as a field enumerator in this study, his tireless support and effort in collecting the data with us and his good humor. Sadly, Ibrahim passed away during the course of this study. He would have met the criteria for authorship on this paper had he lived to see its completion, and we gratefully honor his contribution here.

## List of abbreviations

KAP: Knowledge, attitudes, and practices
MDA: Mass drug administration
ODK: Open Data Kit
WHO: World Health Organization
ZEST: Zanzibar Elimination of Schistosomiasis Transmission

## References

1. World Health Organization. Ending the neglect to attain the sustainable development goals: a road map for neglected tropical diseases 2021–2030. World Health Organization; 2020. Available from: https://apps.who.int/iris/handle/10665/338565.

2. World Health Organization. Weekly epidemiological record. Schistosomiasis and soil-transmitted helminthiases: progress report, 2021. Geneva: 2022 48.

3. World Health Organization. 34th meeting of the International Task Force for Disease Eradication, 19-20 September 2022. 2022 Contract No.: WER No 04, 2023, 98.

4. Efared B, Bako ABA, Idrissa B, Alhousseini D, Boureima HS, Sodé HC, Nouhou H. Urinary bladder *Schistosoma haematobium*-related squamous cell carcinoma: a report of two fatal cases and literature review. Tropical Diseases, Travel Medicine and Vaccines. 2022;8(1):3. doi: 10.1186/s40794-022-00161-x.

5. Colley DG, Bustinduy AL, Secor WE, King CH. Human schistosomiasis. Lancet. 2014;383(9936):2253–64. Epub 2014/04/05. doi: 10.1016/S0140-6736(13)61949-2. PubMed PMID: 24698483.

6. World Health Organization. WHO guideline on control and elimination of human schistosomiasis. Geneva. 2022. Available from: https://apps.who.int/iris/handle/10665/351856.

7. Turner HC, Truscott JE, Bettis AA, Farrell SH, Deol AK, Whitton JM, Fleming FM, Anderson RM. Evaluating the variation in the projected benefit of community-wide mass treatment for schistosomiasis: Implications for future economic evaluations. Parasit Vectors. 2017;10(1):213. doi: 10.1186/s13071-017-2141-5.

8. Toor J, Turner HC, Truscott JE, Werkman M, Phillips AE, Alsallaq R, Medley GF, King CH, Anderson RM. The design of schistosomiasis monitoring and evaluation programmes: The importance of collecting adult data to inform treatment strategies for Schistosoma mansoni. PLoS Negl Trop Dis. 2018;12(10):e0006717. Epub 2018/10/09. doi: 10.1371/journal.pntd.0006717. PubMed PMID: 30296257.

9. Global Schistosomiasis Alliance. Behaviour Change [cited 2026 14.10.2026]. Available from: https://www.eliminateschisto.org/resources/behaviour-change-collection.

10. Knopp S, Ame SM, Person B, Hattendorf J, Rabone M, Juma S, Muhsin J, Khamis IS, Hollenberg E, Mohammed KA, Kabole F, Ali SM, Rollinson D. A 5-year intervention study on elimination of urogenital schistosomiasis in Zanzibar: Parasitological results of annual cross-sectional surveys. PLoS Negl Trop Dis. 2019;13(5). doi: 10.1371/journal.pntd.0007268.

11. Trippler L, Ame SM, Hattendorf J, Juma S, Abubakar S, Ali SM, Kabole F, Rollinson D, Knopp S. Impact of seven years of mass drug administration and recrudescence of *Schistosoma haematobium* infections after one year of treatment gap in Zanzibar: Repeated cross-sectional studies. PLoS Negl Trop Dis. 2021;15(2):e0009127. Epub 2021/02/13. doi: 10.1371/journal.pntd.0009127. PubMed PMID: 33577601.

12. Trippler L, Knopp S, Welsche S, Webster BL, Stothard JR, Blair L, Allan F, Ame SM, Juma S, Kabole F, Ali SM, Rollinson D, Pennance T. The long road to schistosomiasis elimination in Zanzibar: A systematic review covering 100 years of research, interventions and control milestones. Adv Parasitol. 2023. doi: 10.1016/bs.apar.2023.06.001.

13. Stothard JR, Khamis AN, Khamis IS, Neo CHE, Wei I, Rollinson D. Health education and the control of urogenital schistosomiasis: Assessing the impact of the Juma Na Kichocho comic-strip medical booklet in Zanzibar. Journal of Biosocial Science. 2016;48(S1):S40–S55. doi: 10.1017/S0021932016000122.

14. Stothard JR, Mook P, Mgeni AF, Khamis IS, Khamis AN, Rollinson D. Control of urinary schistosomiasis on Zanzibar (Unguja Island): a pilot evaluation of the educational impact of the Juma na Kichocho health booklet within primary schools. Mem Inst Oswaldo Cruz. 2006;101 Suppl 1:119–24. Epub 2007/02/20. doi: 10.1590/s0074-02762006000900019. PubMed PMID: 17308758.

15. Person B, Knopp S, Ali SM, A’Kadir FM, Khamis AN, Ali JN, Lymo JH, Mohammed KA, Rollinson D. Community co-designed schistosomiasis control interventions for school-aged children in Zanzibar. Journal of biosocial science. 2016;48:S56–S73. doi: 10.1017/S0021932016000067.

16. Knopp S, Person B, Ame SM, Ali SM, Mohammed KA, Rollinson D. Elimination of urogenital schistosomiasis in Zanzibar: Challenges for an integrated multidisciplinary research programme. Tropical Medicine and International Health. 2015;20:230–1. doi: 10.1111/tmi.12574.

17. Trippler L, Ali SM, Ali MN, Mohammed UA, Suleiman KR, Ndum NC, Juma S, Ame SM, Kabole F, Hattendorf J, Knopp S. Adaptive integrated intervention approaches for schistosomiasis elimination in Pemba: A 4-year intervention study and focus on hotspots. PLoS Negl Trop Dis. 2025;19(5). doi: 10.1371/journal.pntd.0013079.

18. Ndum NC, Trippler L, Mohammed UA, Ali MN, Hattendorf J, Ame SM, Kabole F, Utzinger J, Ali SM, Knopp S. Effect of behavioural interventions on schistosomiasis-related knowledge, attitudes and practices of schoolchildren in Pemba, Tanzania: Results of a 3-year study. 2025. doi: 10.1371/journal.pntd.0013462.

19. Trippler L, Ali SM, Ame SM, Hattendorf J, Suleiman KR, Ali MN, Juma S, Kabole F, Knopp S. Fine-scale-mapping of *Schistosoma haematobium* infections at the school and community levels and intermediate host snail abundance in the north of Pemba Island: Baseline cross-sectional survey findings before the onset of a 3-year intervention study. Parasit Vectors. 2022;15(1):292. Epub 2022/08/17. doi: 10.1186/s13071-022-05404-6. PubMed PMID: 35974353.

20. Ministry of Finance and Planning - National Bureau of Statistics Tanzania, President’s Office - Finance and Planning - Office of the Chief Government Statistician Zanzibar. Administrative units population distribution report. www.nbs.go.tz/index.php/en/. 2022. Available from: www.nbs.go.tz/index.php/en/.

21. Trippler L, Hattendorf J, Ali SM, Ame SM, Juma S, Kabole F, Knopp S. Novel tools and strategies for breaking schistosomiasis transmission: Study protocol for an intervention study. BMC Infect Dis. 2021;21(1):1024. Epub 2021/10/02. doi: 10.1186/s12879-021-06620-8. PubMed PMID: 34592960.

22. Trippler L, Ali MN, Ame SM, Ali SM, Kabole F, Hattendorf J, Knopp S. GPS-based fine-scale mapping surveys for schistosomiasis assessment: A practical introduction and documentation of field implementation. Infect Dis Poverty. 2022;11(1):8. Epub 2022/01/17. doi: 10.1186/s40249-021-00928-y. PubMed PMID: 35033202.

23. Rassi C, Martin S, Graham K, de Cola MA, Christiansen-Jucht C, Smith LE, Jive E, Phillips AE, Newell JN, Massangaie M. Knowledge, attitudes and practices with regard to schistosomiasis prevention and control: Two cross-sectional household surveys before and after a Community Dialogue intervention in Nampula province, Mozambique. PLoS Negl Trop Dis. 2019;13(2):e0007138. Epub 2019/02/08. doi: 10.1371/journal.pntd.0007138. PubMed PMID: 30730881.

24. Favre TC, Massara CL, Beck LCNH, Cabello RKSA, Pieri OS. Adherence to diagnosis followed by selective treatment of schistosomiasis mansoni and related knowledge among schoolchildren in an endemic area of Minas Gerais, Brazil, prior to and after the implementation of educational actions. Parasite Epidemiology and Control. 2021;13:e00208. doi: 10.1016/j.parepi.2021.e00208.

25. Ayabina DV, Clark J, Bayley H, Lamberton PHL, Toor J, Hollingsworth TD. Gender-related differences in prevalence, intensity and associated risk factors of *Schistosoma* infections in Africa: A systematic review and meta-analysis. PLoS Negl Trop Dis. 2021;15(11):e0009083. Epub 2021/11/18. doi: 10.1371/journal.pntd.0009083. PubMed PMID: 34788280.

26. Adam I, A. AN, Al-Wutayd O, Khamis AH. Prevalence of schistosomiasis and its association with anemia among pregnant women: a systematic review and meta-analysis. Parasit Vectors. 2021;14(1):133. Epub 2021/03/04. doi: 10.1186/s13071-021-04642-4. PubMed PMID: 33653391.

27. Salawu OT, Odaibo AB. Maternal schistosomiasis: A growing concern in sub-Saharan Africa. Pathog Glob Health. 2014;108(6):263–70. Epub 2014/09/17. doi: 10.1179/2047773214y.0000000150. PubMed PMID: 25223633.

28. Young MF. Maternal anaemia and risk of mortality: a call for action. Lancet Glob Health. 2018;6(5):e479–e80. Epub 2018/03/25. doi: 10.1016/s2214-109x(18)30185-2. PubMed PMID: 29571593.

29. Rahmati S, Delpishe A, Azami M, Hafezi Ahmadi MR, Sayehmiri K. Maternal anemia during pregnancy and infant low birth weight: A systematic review and meta-analysis. Int J Reprod Biomed. 2017;15(3):125–34. Epub 2017/06/06. doi: 10.29252/ijrm.15.3.125. PubMed PMID: 28580444.

30. Wolmarans CT, de Kock KN. The influence of health education on the prevalence, intensity and morbidity of *Schistosoma haematobium* infections in children over a two-year period in the Limpopo Province, South Africa. Southern African Journal of Epidemiology and Infection. 2009;24(1):13–7. doi: 10.1080/10158782.2009.11441334.

31. Kokaliaris C, Garba A, Matuska M, Bronzan RN, Colley DG, Dorkenoo AM, Ekpo UF, Fleming FM, French MD, Kabore A, Mbonigaba JB, Midzi N, Mwinzi PNM, N’Goran EK, Polo MR, Sacko M, Tchuem Tchuenté L-A, Tukahebwa EM, Uvon PA, Yang G, Wiesner L, Zhang Y, Utzinger J, Vounatsou P. Effect of preventive chemotherapy with praziquantel on schistosomiasis among school-aged children in sub-Saharan Africa: A spatiotemporal modelling study. The Lancet Infectious Diseases. 2022;22(1):136–49. doi: 10.1016/S1473-3099(21)00090-6.

32. Maddren R, Phillips A, Ower A, Landeryou T, Mengistu B, Anjulo U, Firdawek E, Negussu N, Anderson R. Soil-transmitted helminths and schistosome infections in Ethiopia: a systematic review of progress in their control over the past 20 years. Parasit Vectors. 2021;14(1):97. doi: 10.1186/s13071-021-04600-0.

33. Coulibaly JT, Ouattara M, Barda B, Utzinger J, N’Goran EK, Keiser J. A rapid appraisal of factors influencing praziquantel treatment compliance in two communities endemic for schistosomiasis in Côte d’Ivoire. Trop Med Infect Dis. 2018;3(2). Epub 2018/10/03. doi: 10.3390/tropicalmed3020069. PubMed PMID: 30274465.

34. Inobaya MT, Chau TN, Ng SK, MacDougall C, Olveda RM, Tallo VL, Landicho JM, Malacad CM, Aligato MF, Guevarra JB, Ross AG. Mass drug administration and the sustainable control of schistosomiasis: an evaluation of treatment compliance in the rural Philippines. Parasit Vectors. 2018;11(1):441. Epub 2018/08/02. doi: 10.1186/s13071-018-3022-2. PubMed PMID: 30064469.

35. Mwanga JR, Lwambo NJ. Pre- and post-intervention perceptions and water contact behaviour related to schistosomiasis in north-western Tanzania. Acta Trop. 2013;128(2):391–8. Epub 2012/10/13. doi: 10.1016/j.actatropica.2012.09.017. PubMed PMID: 23058736.

36. Anyolitho MK, Poels K, Huyse T, Tumusiime J, Mugabi F, Tolo CU, Masquillier C, Nyakato VN. Knowledge, attitudes, and practices regarding schistosomiasis infection and prevention: A mixed- methods study among endemic communities of western Uganda. PLoS Negl Trop Dis. 2022;16(2):e0010190. Epub 2022/02/24. doi: 10.1371/journal.pntd.0010190. PubMed PMID: 35196328.

